# “They feel like they have found a friend, and they are able to open-up and talk more”: A qualitative study on the role of Expert Clients and Health Promotion Officers in providing ART adherence support at Lighthouse Trust Clinics in Malawi

**DOI:** 10.64898/2026.08.05.26359835

**Authors:** Odala Sande, Christine Kiruthu-Kamamia, Agness Thawani, Jacqueline Huwa, Gillian O’Bryan, Geldert Davie Chiwaya, Petros Tembo, Vivian Chipolombwe, Hannock Tweya, Lisa Orii, Caryl Feldacker

**Author notes:** Corresponding author, International Training & Education Center for Health (I-TECH) Department of Global Health, University of Washington, Box 359931, Seattle, USA. **E-mail addresses of authors:** OS, CK, AT, JH, GC, PT, VC, GO, HT, LO, CF.

## Abstract

**Introduction:** Expert clients (ECs) and Health Promotion Officers (HPOs) play a similar, and crucial, role in promoting client engagement and retention in antiretroviral therapy (ART) care. At Lighthouse Trust’s large public clinics in Lilongwe, Malawi, ECs/HPOs are HIV-positive clients who provide ongoing counseling to client peers during the first 12 months on ART. This study aims to explore the challenges EC/HPO face to gain insights for improving retention support services.

**Methods:** Using a rapid qualitative study design, ten key informant interviews (KIIs) were conducted with ECs/HPOs at Lighthouse Trust’s two urban clinics. The interviews focused on retention challenges, strategies to increase client retention, and specific recommendations to improve client engagement.

**Results:** Across the KIIs, ECs/HPOs expressed a strong sense of responsibility in supporting and motivating other people living with HIV. ECs/HPOs navigate complex client relationships by building trust and respecting cultural sensitivities. They consider themselves role models for ART adherence, disclosing their own HIV status to foster openness and encourage treatment continuity. While EC/HPOs tailor their conversations to clients’ needs, they remain disappointed by their inability to retain everyone in care. External factors, such as stigma and clients’ Socioeconomic status complicates their efforts. ECs/HPOs noted that ART clients who present as new clients for HIV testing to restart treatment instead of disclosing their treatment gap is particularly difficult to manage. To improve retention, ECs/HPOs recommended non-judgmental re-engagement strategies to encourage lost-to-follow-up clients to return to care, reduced documentation burdens, and promoted broadcasted ART retention messages on radio and television to support their work.

**Conclusions:** The findings highlight the role of ECs/HPOs in building trust and adapting counseling approaches to support client engagement in ART care. Efforts are needed to encourage clients to return to ART services even after treatment gaps, with both EC/HPOs and the broader clinic community fostering a supportive care environment.

## Introduction

Despite global attention, funding, and consistent efforts, retaining people living with HIV (PLHIV) on antiretroviral therapy (ART) in sub-Saharan Africa remains a challenge. Socioeconomic factors such as poverty and stigma impede continuous retention, while health system constraints - like limited healthcare infrastructure and provider shortages - further exacerbate the issue[1–3]. In low- and middle-income countries (LMICs) implementing universal test and treat, nearly 29% of PLHIV disengage from care within 36 months of starting ART[4]. Disengagement from ART care diminishes the immunological benefits of ART and increases risk of AIDS-related morbidity and mortality [5,6].

Improving retention on ART requires targeted interventions addressing both structural barriers and individual needs [7]. At the individual level, HIV programs commonly implement retention strategies that include active patient tracing, community-based ART services, conditional incentives, and peer-support [8]. Peer support, where PLHIV who are retained in ART care provide guidance, encouragement and assistance to early ART initiates has over a decade of evidence that suggests its central role to =improve ART retention. [8,9]

In Malawi, Lighthouse Trust (LT) operates public ART clinics in Lilongwe, offering free, comprehensive HIV care and treatment to approximately 90,000 clients on ART. To improve retention, LT facilities implement Ministry of Health-defined interventions, including active patient tracing, community-based ART services, and peer support.[10] Expert Clients (ECs) and a closely related healthcare cadre, Health Promotion Officers (HPOs), both provide peer support to clients. While ECs and HPOs have similar retention support duties, education requirements differ: ECs require a junior secondary education (JCE) level. In contrast, HPOs are required to have completed secondary education and obtain a Malawi School Certificate of Education (MSCE). All ECs and HPOs undergo training in health education, initial ART assessment and counseling, psychosocial support, as well as phone tracing. ECs are PLHIV who leverage personal experiences to foster peer-to-peer support relationships. HPOs - often also PLHIV-supervise ECs, combining lived experience with more formal health education and counseling training. Each HPO/EC is assigned up to 15 new ART initiates per year, providing ongoing counseling and appointment reminder calls throughout the initial 12-15 months of care. ECs/HPO are employed by LT.

Even though LT has a well-received, multi-prong, differentiated approach to clinic- and community-based care [11] and provides in-person and text-based retention support to aid retention and reengagement in care [12], reaching retention goals is still challenging. Previous qualitative studies on client retention at LT facilities reported financial constraints, such as travel, as reasons for ART disengagement. [13] A recent quantitative study on patient engagement and re-engagement in long-term ART at LT found that a little over half of all clients were retained at 24 months, with higher retention among clients with phones and those enrolled in text-based retention efforts; however, ART retention varied over time and requires differentiated approaches to reach distinct client groups [14]. Therefore, given persistent ART retention gaps at LT and across the region, where 12-month ART retention was 81% in 2023 [15], insights from by ECs/HPOs (also referred to as, “Buddies”), could inform new strategies to increase ART retention. We, therefore, conducted key informant interviews (KII) with EC/HPO Buddies at LT facilities to explore retention challenges and identify potential interventions. Drawing on their deep understanding of daily HIV care realities, the findings of this study aim to enhance retention support at LT and beyond.

## Methods

### Study setting

This rapid qualitative study was conducted at two LT-operated facilities in Lilongwe, Malawi: Lighthouse Clinic (LH) and Martin Preuss Center Clinic (MPC). Combined, the two ART clinics provide free ART services to approximately 38,000 clients.

Individuals with a history of ART but without documented evidence of prior use are retested for HIV. All individuals who test HIV positive, including those newly diagnosed, receive post-test counseling and are assigned to an EC or HPO for further support. (**Fig 1**). The ECs/HPOs document the client’s details, visit date and next appointment date in the ART appointment register. Clients are initiated on ART on the same day. The ECs/HPs escort newly diagnosed HIV individuals through the ART clinic workstations. During this time, ECs/HPOs provide one-on-one counseling on the importance of taking ARVs as prescribed by the ART providers, adherence to clinic appointments, and observing the routine viral load milestone. ECs/HPOs also share their personal experiences in living positively with HIV and adhering to ART, aiming to connect and motivate their clients to do the same.

**Fig 1:**
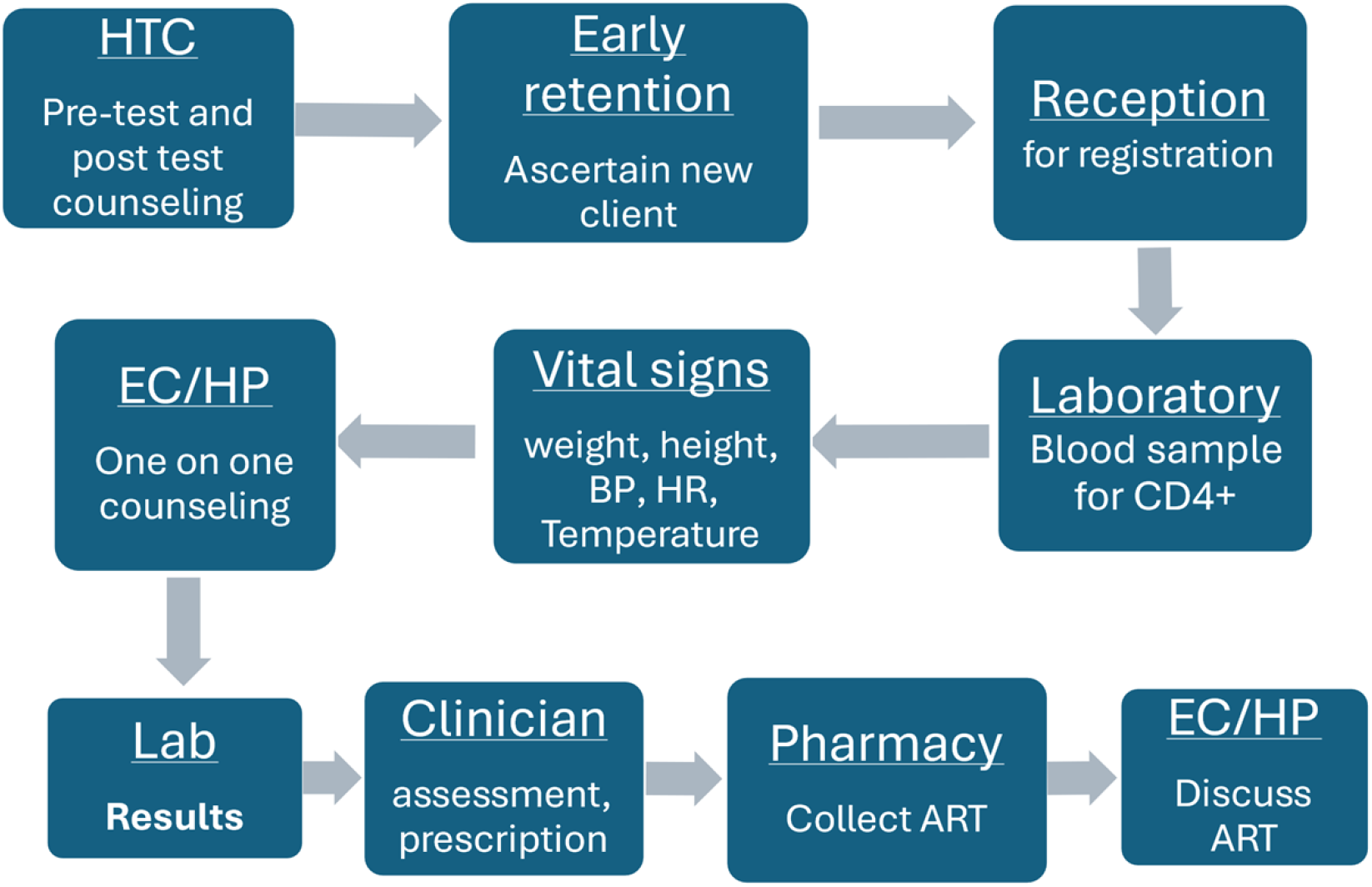
Clinic flow for PLHIV at ART initiation at Lighthouse and Martin Preuss Center, Lilongwe, Malawi.

After ART initiation, ECs/HPOs document the client’s details, visit date and next appointment date in their EC/HPO ART appointment register. A locator form is also completed, capturing the client’s home physical address, personal phone number and the phone numbers of the guardian to facilitate follow-up if the client misses an appointment.

For clients with a documented phone number, ECs and HPOs remind them about the upcoming ART clinic visits via phone calls. If a client misses an appointment, up to 5 attempts are made. Clients who do not return within 14 days after a missed appointment are referred to the Back to Care tracing program which conducts home-based follow-up using the recorded physical address.

### Study population and Data collection

LH and MPC have a total of 10 ECs (5 at each facility) and 8 HPOs (1 at LH and 7 at MPC) who operate similarly across clinics. We interviewed all ECs and HPOs who were available during the data collection periods at both clinics. This approach is intended to capture the full range of frontline retention perspectives at both facilities. Interviews were conducted between 19th June to 26 July, 2024, in private settings by a trained research assistant experienced in qualitative interviews. Each interview lasted approximately 20-45 minutes and was conducted in Chichewa. KIIs were audio recorded, transcribed, translated to English, and de-identified before being shared with the qualitative analysis team.

### Qualitative Analysis

Two investigators deeply familiar with the LT context conducted rapid qualitative analysis of the KIIs. Rapid qualitative analysis [RQA] is an implementation science method to generate rigorous, qualitative results in a shorter time frame compared to traditional qualitative methods and with the aim of informing practice, policy, or program adaptations in real-world settings [16]. RQA was successfully used in a previous qualitative study at LT by this team [12], centering the analysis and results generation firmly within the local setting. In accordance with RQA [16], a discussion guide and transcript summary template was created to guide the KIIs and the analysis [S1 Appendix]. Each investigator then summarized the transcript by topic (“domain”) using bullet points and illustrative quotes to capture the essence of the transcript. Themes and sub-themes were compiled and discrepancies between the two investigators resolved through discussion with the broader study team when necessary. Within the Buddy support activities examined in this study, ECs and HPOs perform the same role; HPOs’ additional supervisory and formal counseling duties lie outside the scope of the activities described here. As the differences in responses between ECs and HPOs were minimal in this small study, and saturation was achieved across frontline workers, the findings were not disaggregated by cadre. The study team reviewed the findings to ensure they reflected the perspectives of the participating ECs and HPOs

### Researcher Reflexivity and Inclusivity in Global Research

The interviewers and analysts are experienced researchers employed by LT but did not routinely interact with or supervise the ECs/HPOs. Their shared institutional affiliation and familiarity with the clinic context likely fostered participant comfort and candor. However, it is also possible that social desirability bias could have led the research team to be more supportive of EC/HPO colleagues or led EC/HPO respondents to be more conservative or favorable in their responses. The author group also includes international researchers based at a U.S. academic institution alongside local researchers embedded in the LT program, and all authors engaged in an iterative process of data review and reflection [17], including group discussions of preliminary results and how individual perspectives may have shaped data interpretation. These potential biases cannot be quantified but were actively considered through this reflective process during analysis and manuscript preparation. Additional information regarding the ethical, cultural, and scientific considerations specific to inclusivity in global research is included in the Supporting Information (S2 Checklist).

### Ethics considerations

The two-way texting (2wT) study protocol to improve client adherence, of which this sub-study was embedded, was reviewed and approved by the Malawi National Health Sciences Research Committee (#20/06/2565) and the University of Washington ethics review board (STUDY000101060). Written informed consent was obtained from all ECs and HPOs prior to participation in the study. Participants were informed that their decision to participate would not affect their employment status.

## Results

### Demographic characteristics of study participants

A total of 10 KIIs were conducted: 5 with ECs (1 from LH and 4 from MPC) and 5 with HPOs, all from MPC. The gender distribution was 4 males, and 6 females. The KIIs revealed several themes concerning EC/HPO [Buddy] support in enhancing client retention, barriers to buddy support intervention and broader intervention suggestions for improving buddy support.

### Drivers of ECs/HPOs efforts

#### Peer identity and commitment to client success

ECs/HPOs are motivated by a strong sense of responsibility and personal commitment to support peers living with HIV, particularly in helping them accept their diagnosis and adhere to ARVs. Drawing on their own experiences, ECs/HPOs said they disclose their HIV status to serve as role models, reassure clients that they are not alone, and that living a healthy, HIV-positive life is possible. As one participant explained: “*We tell them our status so that they should be encouraged by seeing us in case they were disappointed/sad*.” This act of disclosure not only inspires but also motivates newly diagnosed individuals.

Another participant said, *“They should also know that they are not the only ones; there are a lot of us who are HIV positive… that also helps the person to accept it early and start taking medication.”*

Since retaining clients on ART is their responsibility, ECs/HPOs take client loss to follow-up personally, often internalizing blame. As one noted: *“When we see that our clients are not reporting back, it makes us very sad,” and that “it pains us to see someone that we are talking to has turned into a defaulter.”*

#### Facilitating a supportive environment for ART engagement

EC/HPOs believe that their Buddy support services contribute meaningfully to client retention by reminding clients of their appointment dates via phone calls, thereby helping them adhere to their appointment dates. In the clinic, ECs/HPOs accompany their clients throughout the clinic flow, ensuring that clients learn the clinic routine.

> *“We keep on encouraging them to make sure that we should help them at every point where they are supposed to reach [at the clinic], whether they must go to the Lab, or have their vitals checked or to the doctor to get the drugs, we are supposed to help them on everything during their first visit.”*

In addition to these reminders, ECs/HPOs provide ongoing counseling to assist clients in navigating the challenges associated with ART use. The buddies create an environment where clients are free to share their anxieties and experiences on ART.

> “*There is no one who they can open-up to but when they get to us, they feel like they have found a friend and they are able to open-up and talk more, as well as the challenges that they face since they started taking the drugs”*

By providing ongoing support, following up on medication adherence and missed appointments, *“we make a good friendship with the person.”* This connection helps clients feel understood and supported, reducing anxiety and instilling hope.

### Barriers to Buddy support

All peer supporters noted that not all clients could be retained in care. This led them to fear that their lower-than-expected retention rates may reflect poorly on their performance as ECs/HPOs, adding pressure to an already challenging job. Several themes emerged on potential weaknesses within the Buddy program.

#### Disclosure fears among returning ART clients

Across all KIIs, the most frequently noted challenge to retention efforts were non-ART naïve clients seeking HIV testing to re-initiate ART without disclosing their HIV positive status. These clients pose challenges for EC/HPOs due to the largely unnecessary time commitment that it takes to provide initiation services for those who have already received this extension counseling. These “re-initiates” might have moved to the city for work and run out of ARVs, defaulted from ART care and sought to resume ART treatment, or fallen sick and sought medical care.

> “*The other challenge is those clients who seem to be new, yet they are defaulters from other facilities. We can reach at that conclusion because we have been doing this for long time, we have been on medication for long. We have experience. Some of the sickness originates from the client not taking medication.”*

ECs/HPOs reported that some clients preferred to restart ART as a new initiate rather than face other clinicians or HCWs after a long treatment interruption. Fear of being shouted at or poorly treated by staff leads them to undergo the lengthy process of new HIV testing and initiation counseling rather than admitting they were previously on ART. Some clients fear the responses of clinicians if they return after a treatment interruption, noting that *“they were afraid that we will shout at them. We tell them that we do not shout at them and encourage them to be adherent.”* It is known that some clinicians do not want to help initiate new clients due to the burden of starting clients in care.

> *“Sometimes when we have escorted the initial client to the clinician (not all clinicians), but it may happen that they would refer again to the next-door clinician because they just don’t want to handle the client since they do undergo a lot of processes.”*

ECs/HPOs also discussed how these “new ART initiates” may create issues in data quality and in retention statistics. These unconfirmed reinitiates may return to their local clinics without a formal transfer, appearing to default at Lighthouse.

> “*We meet some clients who already started taking the drugs, they may come to the facility and lie that have not been initiated…They get tested as a new client when they report to the clinic. We end up having defaulters when such people sell their goods and go back home* [outside Lilongwe]*”*.

#### Lack of mobile phone access or wrong mobile numbers

Several client issues create retention challenges. First, some retention efforts require clients to have phones, but some clients lack phones or lack access to phones. ECs/HPOs thought that some wrong information was due to a lack of disclosure or persistent stigma in the community or family, which discourages clients from accepting follow-up by phone calls or home visits. Additionally, the lack of verifiable contact information complicated follow-up efforts.

> “*For clients who do not have phones, it is very difficult because when we leave each other today we will meet again … when they report to the clinic, it is easy for them to forget or not report to the clinic. That makes us very sad, ‘did I not chat with them properly?’ or what happened?”*

Second, some clients provide inaccurate phone numbers or addresses for follow-up or tracing. This impedes communication, leaving the retention team unable to help retain a client until they present themselves at the clinic.

> “*That is one of the things that is challenging because a lot of people give wrong numbers, even the maps* [to their homes] *that they give us are not true and they know that they will not report to the clinic.”*

ECs/HPOs thought that some wrong information was due to lack of disclosure or persistent stigma in the community or family which discourages clients from accepting follow-up by phone calls or home visits. Additionally, lack of verifiable contact information complicated follow-up efforts.

All peer supporters noted that not all clients could be retained in care. This led them to fear that their lower-than-expected retention rates may reflect poorly on their performance as ECs/HPOs, adding pressure to an already challenging job.

#### Clients, especially those with greater challenges, require retention support differentiation

Not all clients present the same retention challenges, requiring differentiated support. Some clients need special attention. New initiates can be harder to counsel as, *“There are a lot of challenges within three months unlike after we know the person has settled”.* Also, clients who have other sexually transmitted infections (STIs) at ART initiation are difficult to retain on ART care because they often believe ART is no longer needed when other STI is cured.

> *“It is possible for things not to go well, some people…maybe they want to get help for STIs. These are the people who are difficult because once their problem is gone, it is difficult for these people to report back* [for ART visits]*”*.

Additionally young people may also require special attention, as they are not mature and may find it difficult to accept their status.

> “*Age group 25 below, this is one of the groups that think twice before doing something, they are different from someone who is mature. This group needs special treatment. We are supposed to give them information that is very comprehensive so that when they go home, they should know that we are thinking about them”*

### Improvements to enhance ART retention

#### Provide phones to retention support teams

Each ECs/HPOs suggested that they need their own work phone to contact clients and have a clear call-back number for their clients who want to reach them for support. Many also noted that they desired to be acknowledged and recognized by the clinic and the staff for their positive and important role in providing proper support to clients, thereby improving overall retention. This would improve the ECs/HPOs dedication to their work. ECs/HPOs would also like to be recognized by increasing their monthly salaries.

### Reduce the monitoring and evaluation (M&E) workload

ECs/HPOs also noted the need for reduced M&E paperwork for client tracking. One EC lamented that they are supposed to follow-up on their clients in multiple forms, *“which takes me about 2-3 days. I check an individual hardcover of their dates for example this one is going to come in August, then I need to write down on this paper/forms. This is tiresome.”* The need to track clients manually or via redundant, overlapping monitoring tools was considered overwhelming and unnecessary, diverting their attention from direct patient care to administrative tasks.

> *“I feel like there is a lot of paperwork at early retention* [EC/HPO worksite]*. As such it is one of the things that I see to be making the treatment buddy weak. We have a lot of documentation. One client is documented in several places for the same issues. The whole day will go by spent on writing in the daily retention, forgetting to make the phone calls, reminders, follow up for the people who are coming because I am busy writing the papers…With daily monitoring, we forget the actual work on the ground.”*

#### Consider additional media interventions

ECs/HPOs suggested improvements in retention support including development of an HIV education and treatment management app for phones and disseminating ART-related information via radio, phones, and televisions.

> *“I was thinking that, if possible, anyone who is starting ART at the facility and wants information at once can just sign in to the app and they will be able to find anything that they want. Let’s say I am at home, I have an ART number and would want to know the next appointment date, if we have that application, one can just punch in the ART number and will get all their information. Maybe because they say that clinic stuff shouldn’t be everywhere as part of security, but I feel like it can be a good way for the person to follow their health, even about viral load.”*

The utilization of radios and television programs to disseminate ART information can help to reach PLHIV, on ART and not, and others whose awareness of HIV may have faded over time.

> *“Maybe there is no radio program that encourages people to return to get medication or the benefit of getting medication (refill). If there can be an initiative through a radio program, the radio reaches a lot of people and that can help us to have high retention rates as they will talk frequently about the benefit of taking medication.”*

With largely free access to both radio and national TV, the opportunity to produce short programs or public service announcements could provide a vital new channel to increase awareness of retention problems and clinic openness for clients who want to return to care but are concerned about their clinic reception.

## Discussion

In this qualitative study, we gained a more nuanced perspective of how ART retention officers retain ART clients and the challenges they face. Our findings revealed that ECs and HPOs employ continuous, adaptive approaches to engage clients in care, tailoring their methods to clients’ individual needs to optimize treatment adherence. These peer supporters demonstrate flexibility in supporting ART clients, prioritizing client-centered interactions over standardized protocols to foster personal connections and motivate sustained ART adherence. Reflecting upon their insights into client-centered approaches to enhance HIV retention, we share several lessons learned on engaging clients in care from this small group of peer PLHIV experts.

First, ECs/HPOs believed creating rapport through an informal, empathetic, conversational style and interactive, client-led discussion was critical. This style aligns with principles of motivational interviewing (MI), supporting clients to remain in care by encouraging behavior change and fostering acceptance [18,19]. This approach may also build trust swiftly through shared experience, promoting a sense of community and enabling open dialogue about real-world challenges related to lifelong ART adherence. In an MI study in South Africa, MI participants were more compliant with appointments and were more likely to achieve viral load suppression as compared to peers who did not receive MI (65.3% vs. 49.3% in controls), and they reported more positive counseling experiences and greater confidence in managing their treatment [20]. At LT, the empathetic approach helped clients accept their HIV status and empowered them to set personal goals and practical strategies for adherence, encouraging new ART clients to be accountable for their health outcomes.

Second, across the board, ECs/HPOs had challenges with non-ART naïve clients who presented for HIV testing to re-initiate ART without disclosing their HIV positive status. This adds additional workload and also may artificially reduce retention numbers, which may negatively impact perceptions of EC/HPO service quality. A study from South Africa to explore why some PLHIV on ART may retest found that some clients who return to retest may want confirmation of their status; clients may not understand that the medications they currently take are ART; or they want to be retested in a clinic where they are confident in receiving quality care [21]. For some, retesting for HIV status may signal that they are ready to return to ART after an interruption [22], that they reached acceptance of their status, or when they wanted to change health facilities [23], potentially not understanding the transfer process. In Malawi, Lighthouse colleagues also suggested that some people on ART may present as new initiates having exhausted their “emergency” ART supplies – a MoH granted right to all ART clients to access a 1-month ART supply in any clinic without transferring care. Clients may need more ART to support travel or work needs, but the process of acquiring more ART is unclear or unknown. Other clients may have defaulted who fear being shouted at by clinic staff for missing visits, driving some to change names or use alternate IDs to appear as new clients to reengage in ART care.

While we did not quantify this phenomenon of retesting in this study, previous research indicates that it is not uncommon for previous ART clients to return to care without disclosing their HIV positive status. In Malawi, the risk ratio of re-testing within the past year among PLHIV aware of their status and on ART compared to PLHIV aware of their HIV status but not on ART is 0.61, ranging from 0.45 to 0.81 [24]. In a large meta-analysis of PLHIV underreporting their true status, the pooled proportion of misreporting was 20%, with results from African studies ranging from 14-52% [25], suggesting that this problem is widespread. While all clients deserve the counseling and care needed to remain on lifelong treatment, the workload impact of additional, unnecessary, HIV testing, initiation counseling, and clinician review, including for ECs/HPOs, is significant, likely reducing scarce resources for other clients and programs. Clients returning to ART care as *new* ART clients if they are not identified by ECs/HPOs, rather than as *defaulters*, also likely skews both clinic and national data on HIV positive testing yield, new HIV infections, ART initiations, and defaulter data. The double-counting of individuals as both new initiates and defaulting also undermines the integrity of treatment programs and compromises the accuracy of tracking treatment outcomes, muddling data needed for resource allocation or treatment distribution decisions. To effectively support clients returning to care, clinics must foster trust, ensuring that the welcoming, non-judgmental environment created by peer supporters extends across all clinic interactions and personnel.

Lastly, despite EC/HPOs efforts and dedication, not all clients can be retained in ART care, risking demotivated Buddies. ECs and HPOs noted that while they provide the same core services to all clients, certain groups—including new initiates and adolescents—require additional support to remain in care. Clients with co-morbidities, alcohol use, mental health conditions, or those with concurrent sexually transmitted infections, face added challenges and require more intensive or integrated services to support effective retention. Persistent community stigma may also discourage some clients from sharing accurate contact information or agreeing to be followed up at home, meaning location information provided to ECs/HPOs may sometimes be intentionally incorrect. These gaps may cause anxiety among ECs/HPOs who fear that sub-optimal retention would reflect poorly on them or be seen as poor quality service provision. While retention efforts are focused on the clients, it is also likely that this critical care cadre would also benefit from additional job retention support. Consideration of additional incentives, acknowledgement, and affirmation of their important role in the overall success of the clinic could further enhance EC/HPO commitment. These findings are also consistent with the Malawi MoH National HIV Strategic Plan [15], which recognizes ECs and other lay health cadres as integral to strengthening community-based support systems and improving ART adherence and retention. Our findings suggest that achieving these national priorities will require continued investment in ECs and HPOs, including reducing administrative burden, providing adequate resources and recognition, and enabling them to focus on delivering client-centered adherence and retention support.[15]

This study had several limitations. First, while the Lighthouse research team took steps to remain objective, their involvement in conducting and analyzing the data may have introduced bias of unknown direction. Second, the qualitative study was not intended to compare ECs and HPOs who complete similar client services; rather, the KIIs encompassed almost all the HCWs in this frontline retention work at LT and other ART clinics. Third, both study sites are urban ART clinics in Lilongwe; findings are likely dissimilar to those that may be found in rural or even lower resourced clinic settings. Perspectives from clients were considered previously and, therefore, not included in this manuscript [11,12]. Lastly, RQA was chosen over traditional thematic analysis given the objective to generate actionable findings for Lighthouse Trust’s retention program, potentially limiting the depth of thematic analysis. Despite these limitations, the retention insights provided by PLHIV who assist in retaining their peers in care is valuable to improving client engagement in ART care.

## Conclusion

Moving forward, there are several suggestions that Lighthouse and other clinics should consider improving the work of frontline retention officers and ART client retention. First, provide tailored support for vulnerable populations such as adolescent and clients with co-morbidities. Secondly, MoH policies should be communicated more effectively to all ART providers and clients to increase awareness of these ART access rights. Thus, creating an enabling and inviting environment for clients seeking temporary care or ART-related medications in other clinics. Lastly, streamlining documentation. processes is essential to ensure that these retention support personnel can focus on their primary role of providing retention support.

## Supporting Information

### S1 Appendix: Interview Guide

Discussion guide and transcript summary template used to conduct and analyze the key informant interviews.

### S2 Checklist: PLOS Inclusivity in Global Research Questionnaire

Completed questionnaire addressing the ethical, cultural, and scientific considerations specific to inclusivity in global research, as required by PLOS ONE policy.

## Authors’ contributions

**Conceptualization**: Hannock Tweya, Caryl Feldacker

**Data curation**: Odala Sande, Christine Kiruthu-Kamamia, Agness Thawani, Jacqueline Huwa, Geldert Chiwaya, Vivian Chipolombwe, Petros Tembo

**Formal Analysis**: Odala Sande, Caryl Feldacker, Gillian O’Bryan, Lisa Orii

**Funding acquisition**: Caryl Feldacker, Hannock Tweya

**Investigation**: Odala Sande, Hannock Tweya, Caryl Feldacker, Jacqueline Huwa, Geldert Chiwaya, Christine Kiruthu-Kamamia

**Methodology**: Odala Sande, Jacqueline Huwa, Hannock Tweya, Caryl Feldacker

**Project administration**: Hannock Tweya, Caryl Feldacker, Jacqueline Huwa, Marrianne M Holec

**Validation**: Gillian O’Bryan, Christine Kiruthu-Kamamia, Lisa Orii, Agness Thawani, Jacqueline Huwa.

**Visualization**: Odala Sande, Hannock Tweya

**Writing-original draft**: Odala Sande, Christine Kiruthu-Kamamia, Vivian Chipolombwe, Hannock Tweya, Caryl Feldacker, Jacqueline Huwa, Agness Thawani, Petros Tembo, Marrianne M Holec

**Writing** – Review & Editing: All authors reviewed and edited

## Data Availability

Data from sensitive interviews about HIV-related services cannot be shared publicly and it protected by heightened privacy standards. De-identified data are available from the Lighthouse Trust for researchers who meet the criteria for access to confidential data. For access to the data via a data sharing agreement and confidentiality agreement, please reach out to Lighthouse IRB officer for this study

## Acknowledgements

The authors would like to thank all the staff who collected data at Lighthouse and Martin Preuss Center clinics. We thank numerous donors supporting the Lighthouse and Martin Preuss Center clinics.

